# Revealing the extent of the COVID-19 pandemic in Kenya based on serological and PCR-test data

**DOI:** 10.1101/2020.09.02.20186817

**Authors:** John Ojal, Samuel P. C. Brand, Vincent Were, Emelda A Okiro, Ivy K Kombe, Caroline Mburu, Rabia Aziza, Morris Ogero, Ambrose Agweyu, George M Warimwe, Sophie Uyoga, Ifedayo M O Adetifa, J Anthony G Scott, Edward Otieno, Lynette I Ochola-Oyier, Charles N Agoti, Kadondi Kasera, Patrick Amoth, Mercy Mwangangi, Rashid Aman, Wangari Ng’ang’a, Benjamin Tsofa, Philip Bejon, Edwine Barasa, Matt. J. Keeling, D. James. Nokes

**Affiliations:** Kenya Medical Research Institute (KEMRI) - Wellcome Trust Research Programme (KWTRP), Kilifi, Kenya; London school of Hygiene and Tropical Medicine (LSHTM), UK; The Zeeman Institute for Systems Biology and Infectious Disease Epidemiology Research (SBIDER), University of Warwick, UK.; School of Life Sciences, University of Warwick, UK.; Health Economics Research Unit, KEMRI-Wellcome Trust Research Programme, Nairobi, Kenya; Population Health Unit, Kenya Medical Research Institute - Wellcome Trust Research Programme, Nairobi, Kenya; Centre for Tropical Medicine and Global Health, Nuffield Department of Medicine, University of Oxford, Oxford, United Kingdom; Department of Infectious Diseases Epidemiology, London School of Hygiene and Tropical Medicine, London, United Kingdom; School of Public Health, Pwani University, Kenya; Ministry of Health, Government of Kenya, Nairobi, Kenya; Presidential Policy & Strategy Unit, The Presidency, Government of Kenya

## Abstract

Policy makers in Africa need robust estimates of the current and future spread of SARS-CoV-2. Data suitable for this purpose are scant. We used national surveillance PCR test, serological survey and mobility data to develop and fit a county-specific transmission model for Kenya. We estimate that the SARS-CoV-2 pandemic peaked before the end of July 2020 in the major urban counties, with 34 - 41% of residents infected, and will peak elsewhere in the country within 2-3 months. Despite this penetration, reported severe cases and deaths are low. Our analysis suggests the COVID-19 disease burden in Kenya may be far less than initially feared. A similar scenario across sub-Saharan Africa would have implications for balancing the consequences of restrictions with those of COVID-19.

## Main Text

The potential risk from SARS-CoV-2 to Africa was identified early in the global pandemic (*1*). As the epicenter of transmission moved from East Asia to West Asia and Europe and then to North America, there was speculation as to the likely impact of the pandemic on the African continent with its young populations, infectious disease burden, undernutrition and fragile health infrastructure. However, as health systems and economies of high-income countries strained, the reported burden of COVID-19 cases and associated deaths in Africa remained low (*2*). Even today, other than for South Africa, this remains true throughout sub-Saharan Africa. The question is whether this is the result of lower risk due to demographic structure (young age (*3*)) or cross-reacting immunity (e.g. pre-existing SARS-CoV-2 cross-reactive T cells (*4*)) a low reproduction number from imposed interventions (such as school closures and lockdowns (*5*)) or environmental conditions (e.g. temperature and humidity (*6*)), or under-reporting. The reason this remains a conundrum is, at least in part, a paucity of good quality data to reveal the probable extent of SARS-CoV-2 spread in African populations.

Following the first confirmed COVID-19 case in Kenya on 13th March 2020, the Kenyan Government moved rapidly, closing international borders, schools, restaurants, bars and nightclubs, banning meetings and social gathering and imposing a dusk to dawn curfew and movement restrictions in the two major city counties, Nairobi and Mombasa, considered epidemic hotspots at the time (*7*). The major concerns from unmitigated spread were a limited surge capacity of the Kenyan health system (*8*) and groups of the Kenyan population identified as potentially highly vulnerable to infection and/or severe disease (*9*). Throughout the months of April, May and into June 2020 few people in Kenya were reported SARS-CoV-2 test positive by polymerase chain reaction (PCR), or severely diseased or dying with COVID-19 as the established cause (*10*). There followed a relaxation of some measures in June and July including opening of restaurants and places of worship and the removal of travel restrictions into and out of Mombasa and Nairobi counties. As of 10^th^ August 2020, there have been 23,873 laboratory-confirmed positive swab tests out of over 320,000 tests, and 391 deaths with a positive test result in Kenya. This should be compared with the 200-250,000 cases and 30-40,000 deaths attributable to SARS-CoV-2 for similar sized countries in Europe (France, Italy, UK) at equivalent months into their epidemics.

The reason for this apparently low level of COVID-19 disease in Kenya is unknown, and non-reporting is a potential explanation. However, two pieces of information suggest an explanation that SARS-CoV-2 has spread extensively with a smaller proportion than expected having severe disease. First, a regionally-stratified seroprevalence study of 3098 Kenyan blood donors sampled between May and June reported a national estimate of 5.2% (adjusted to reflect the population distribution by age, sex and region) (*11*). Seroprevalence was higher in Nairobi (8.5%) and Mombasa (9.3%). These levels of seropositivity are comparable to those reported in May in the UK (*12*), April/May in Spain (*13*), and March/April in some US cities (*14*), where in contrast to Kenya, high numbers of PCR-positive cases, hospitalizations and deaths have also been reported. Second, early in August 2020, we noticed that test-positive PCR cases were declining in Mombasa, the second largest city in Kenya. This went counter to evidence of increased mixing, and hence reproduction potential, arising from Google Mobility data for Mombasa showing a steady reversion in mobility towards pre-COVID-19 intervention levels from early April (Fig. S1).

We developed a simple SEIR compartmental mechanistic and data-driven transmission model for Kenya, which integrates three sources of longitudinal data: national time series PCR tests, the Kenyan serological survey and Google mobility behavioural data (*15*). Our aim is to derive a coherent picture of SARS-COV-2 epidemiology in Kenya and reveal the historic and future patterns of spread across the country and by county. This approach allows us to generate a crude estimate of infection-to-fatality ratio (IFR_crude_) between estimated infections and observed deaths with a positive test result; reported deaths are not used as primary data for inference, but rather as a validation data set for model predictive accuracy (see *supporting information* for description of model validation).

As at 10^th^ August 2020, a substantial proportion of PCR positive tests have been samples from the capital Nairobi (10,575 positive tests), and Kenya’s second largest city, Mombasa, has reported the next highest number of PCR positive tests (1,962). We find that the rate of new infections peaked on May 31st 2020 (CI May 25th - June 15th) in Mombasa and July 10th 2020 (CI July 2nd - July 19th) in Nairobi, and is now declining (Fig. 1H,G). The model suggests that the PCR test and serology data can be explained by the initial presence of < 10 infected individuals in Mombasa and < 50 in Nairobi on 21st February, three weeks before the first reported case in Kenya. Thereafter, growth of transmission was rapid in both counties. In early March, the reproductive ratio was 2.23 and 2.01 in Mombasa and Nairobi, respectively, and the doubling-time estimates were 3.85 and 4.55 days, respectively. After March, the transmission curves flattened substantially. By April 16^th^, the reproductive ratio was 1.18 and 1.04, respectively and the doubling times were 27.7 and 93.5 days, respectively. This change is consistent with the introduction of containment measures by the Kenyan government, and evidence of substantial reduction in mobility (see Google Mobility data Fig. S1). From late April through May and June and into July the evidence suggests movement restrictions became steadily less effective (Fig. 2; Fig. S1). The waning effectiveness of movement restrictions results in an inferred increase in R and an increased rate of epidemic growth; however, in Mombasa and Nairobi we predict that reduction in susceptibility of the population had already caused the effective reproductive ratio (R_eff_; the mean number of secondary cases accounting for reduced susceptibility) to fall significantly below the basic R value (Fig. 2).

**Fig. 1.**
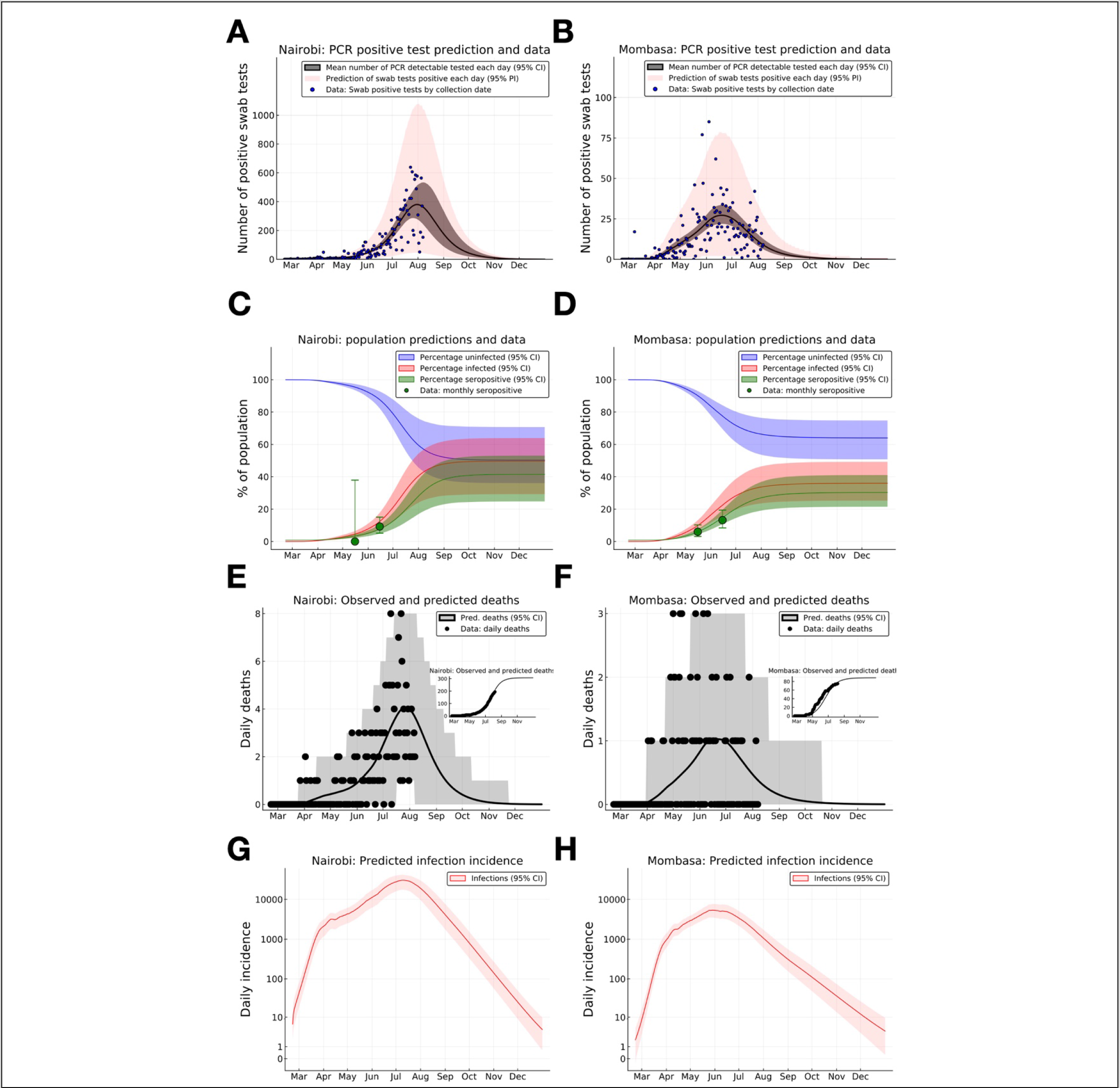
SARS-CoV-2 PCR positive swab tests, seroprevalence and deaths in Nairobi and Mombasa, Kenya, with model forecasting. (**A**) & (**B**) Daily reported positive PCR positive swab tests (*blue dots*) for Nairobi (**A**) and Mombasa (**B**), model prediction of mean daily detection of new PCR-positive swab tests by date of sample collection (*black curve*), and the model prediction interval for observed daily PCR-positive swab tests including inferred day-to-day variation in detection (*pink shading*). (**C**) & (**D**) Monthly seropositivity of Kenya National Blood Transfusion Service (KNBTS) blood donors in Nairobi (**C**) and Mombasa (D) (*green dots*), model predictions for population percentage of seropositivity (*green curve*), exposure to SARS-CoV-2 (*red curve*), and uninfected (*blue curve*). (**E**) & (**F**) Daily deaths with a positive SARS-CoV-2 test in Nairobi (**E**) and Mombasa (**F**) by date of death (*black dots*), and model prediction for daily deaths (*black curve*). Inset plots in (**E**) and (**F**) indicate cumulative reported deaths and model prediction. (**G**) & (**H**) Model estimates for rate of new infections per day in Nairobi (**G**) and Mombasa (**H**). Background shading indicates 95% central credible intervals. Dates for all graphs mark the 1st of each month.

**Fig. 2.**
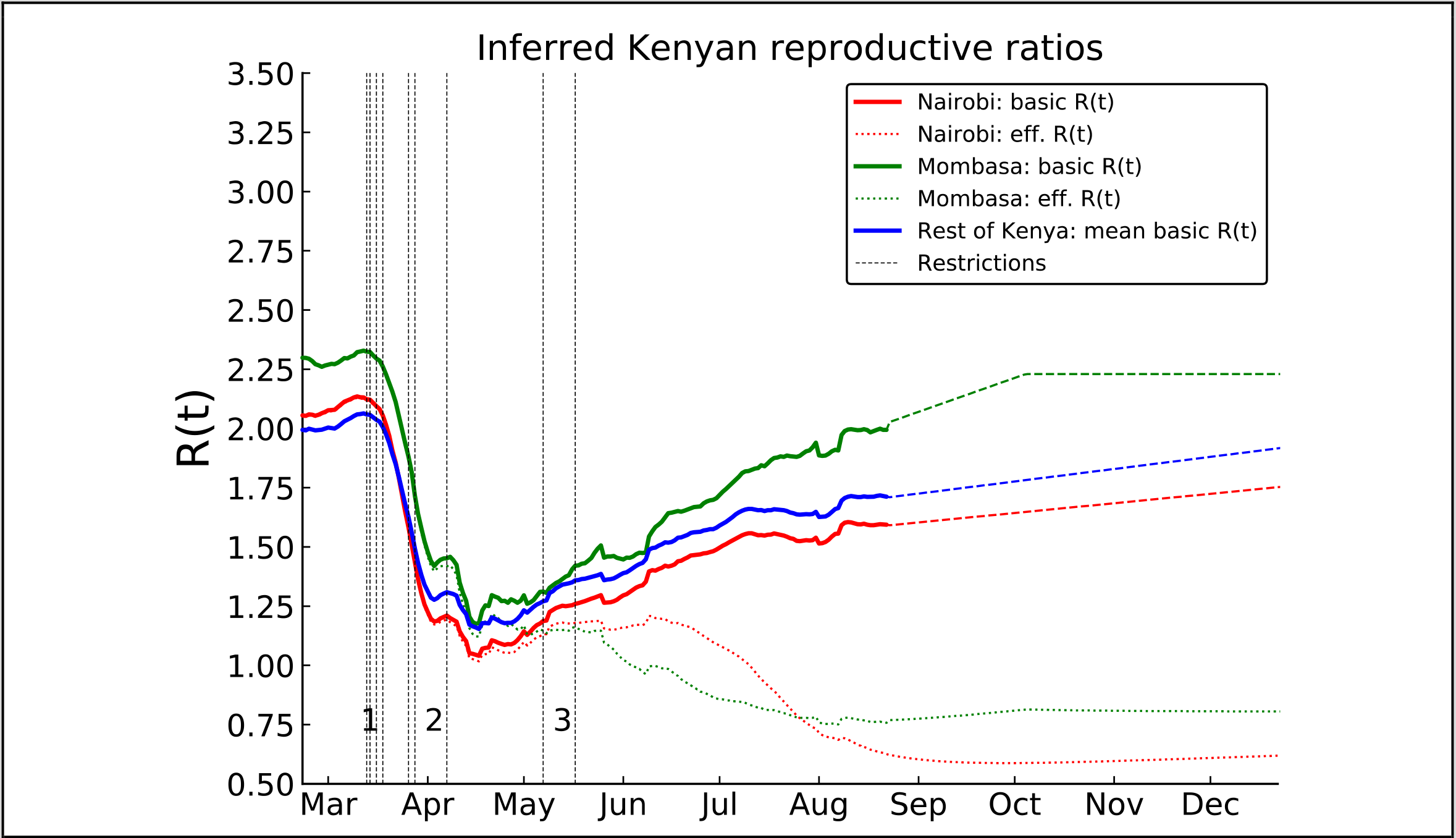
Estimated basic and effective reproductive numbers in Kenya since Feb 21st 2020. The posterior mean reproductive number for Nairobi (*red curves*), Mombasa (*green curves*), and the averaged over reproductive number estimates for all other Kenyan counties (*blue curve*). Shown are both the basic reproductive numbers (expected secondary infections in a susceptible population adjusted for mobility changes since the epidemic start; *solid curves*), their near-term projections (*dashed curves*), and effective reproductive numbers (expected secondary infections accounting for depletion of susceptible prevalence in the population; *dotted curves*). The effective reproductive number varied significantly from county to county and is not shown. Restrictions aimed at reducing mobility in risky transmission settings (*black dotted lines*) are labeled in groups. The chronologically ordered restrictions in each group are: 1) First PCR-confirmed case in Kenya, suspension of all public gatherings, closure of all schools and universities, and retroactive quarantine measures for recent returnees from foreign travel, 2) suspension of all inbound flights for foreign nationals, imposition of a national curfew, and regional lockdowns of Kilifi, Kwale, Mombasa and Nairobi counties, and 3) additional no-movement restriction of worst affected areas within Mombasa and Nairobi, and, closure of the border with Somalia and Tanzania.

By accounting for the delay of an average of 19 days between infection and death (*supporting information* for details on infection to death distribution) we find the transmission curve, estimated from PCR tests and serology, generates a good prediction of the observed daily deaths in Nairobi and Mombasa (Fig. 1 E&F). We emphasize that we did not use daily death data in transmission model inference, therefore the good fit to the observed trend in deaths with a PCR-confirmed test result represents an out-of-sample validation of the modelling (*16*). We predict the peak of positive PCR test samples occurred at the end of July or early August in Nairobi and earlier, mid-June, in Mombasa. The lag between transmission peak and positive swab testing peak being explained by both the delay between infection and becoming detectable by PCR, and the period after an infected individual has ceased being actively infectious but remains detectable by PCR (*17*) (Fig. 1 A&B). As of the beginning of August 2020 we estimate that about 28.5% (CI 17.3%-38.9%) of the Nairobi population, and 27.1% (CI 19.2-37%) of the Mombasa population would be serologically positive with SARS-CoV-2, (Fig. 1 C&D). This estimated level of seropositivity is substantially higher than has been estimated in countries that have been hit hard by the pandemic (*12*-*14*) however they are in broad agreement with a recent study in Niger state, Nigeria (*18*). We caution over-interpreting the forecasts of seroprevalence; the projections could be markedly lower if we assumed significant decay in specific antibody over the timescale of observation of a few months. Importantly, our projections of the proportion exposed are not conditional on the assumption of waning seropositivity.

Accounting for the sensitivity of the serological assay, and the delay between infection and seroconversion, we estimate that the actual exposure of the population to SARS-CoV-2 by August 1st was 40.9% (CI 24.3%-54.7%) in Nairobi and 33.8% (CI 23.7%-46.5%) in Mombasa (Fig. 1 C&D). Such levels of population exposure are predicted to be associated with herd-immunity in these urban populations given the estimated reproductive numbers and effective population size at risk of exposure (P_eff_) due to heterogeneity in the susceptibility, transmissibility and social interactivity in the population (*supporting information* for more details on effective population size in transmission modelling); the effective population size for Nairobi was inferred as 77.0% of actual population size (CI 45.4%-98.8%), for Mombasa 56.4% (CI 39.6%-76.8%). These low herd immunity estimates rest upon inferred variation in risk across the population and reduced reproductive potential due to present interventions. There remains a possibility of future increase in transmission if population mobility continues to rise, if population mixing patterns alter leading to changed risk heterogeneity or if immunity is short lived, leading to a rebound in reported cases. It is possible that non-antibody mechanisms of immunity (for instance T cells (*3*)) also contribute to apparent herd immunity at low serological prevalence.

The fitted IFR_crude_ values for both Nairobi (IFR_crude_ = 0.014% (CI: 0.010%-0.023%)) and Mombasa (IFR_crude_ = 0.02% (CI:0.014%-0.028%)) are substantially lower than the age-adjusted IFR expected for Kenya under full ascertainment (IFR_verity_ = 0.26%; (*19*) and *supporting information*). This is a crude observational value for the infection to fatality ratio, since we do not currently have an estimate of the reporting bias of deaths of individuals infected with SARS-CoV-2. Therefore, our estimate of IFR_crude_ potentially reflects lower detection in Kenya compared to China, as well as any lower mortality risk due to fewer comorbidities.

We extended our model-based inference to each of the 47 counties in Kenya (see dataset S1 for parameter estimates, peak time estimates and IFR_crude_ estimates for each county). For counties with enough data we replicated the inference approach used for Nairobi and Mombasa counties. Model inference for counties with no serological samples and only a small number of PCR positive samples used strong priors inferred from the posterior distributions of model parameters of similar counties which could be fitted (Fig. 3A; *supporting information* for details on how counties were matched to similar counties). We find that, in addition to the two main Kenyan city counties, the rate of new infections in semi-urban counties neighbouring Nairobi (Kiambu, Kajiado, and Machakos) and Mombasa (Kwale), and the Ugandan border county Busia, also peaked before August 2020; Kwale county (neighbouring Mombasa on the Tanzanian border) is predicted to have been the first to peak (posterior mean of peak time May 21^st^ 2020). Note, these early peaks in Kwale and Busia counties might be an artifact of cross-border importations and a testing strategy skewed towards a highly mobile population (i.e. truck drivers). However, the infection rate is predicted to still be rising in other more rural counties, most notably in counties constituting the former Rift valley, Eastern and North Eastern provinces. We forecast that the final county to reach peak infection rate will be the arid and sparsely populated Wajir county (posterior mean 24th September 2020; Fig. 3A).

**Fig. 3.**
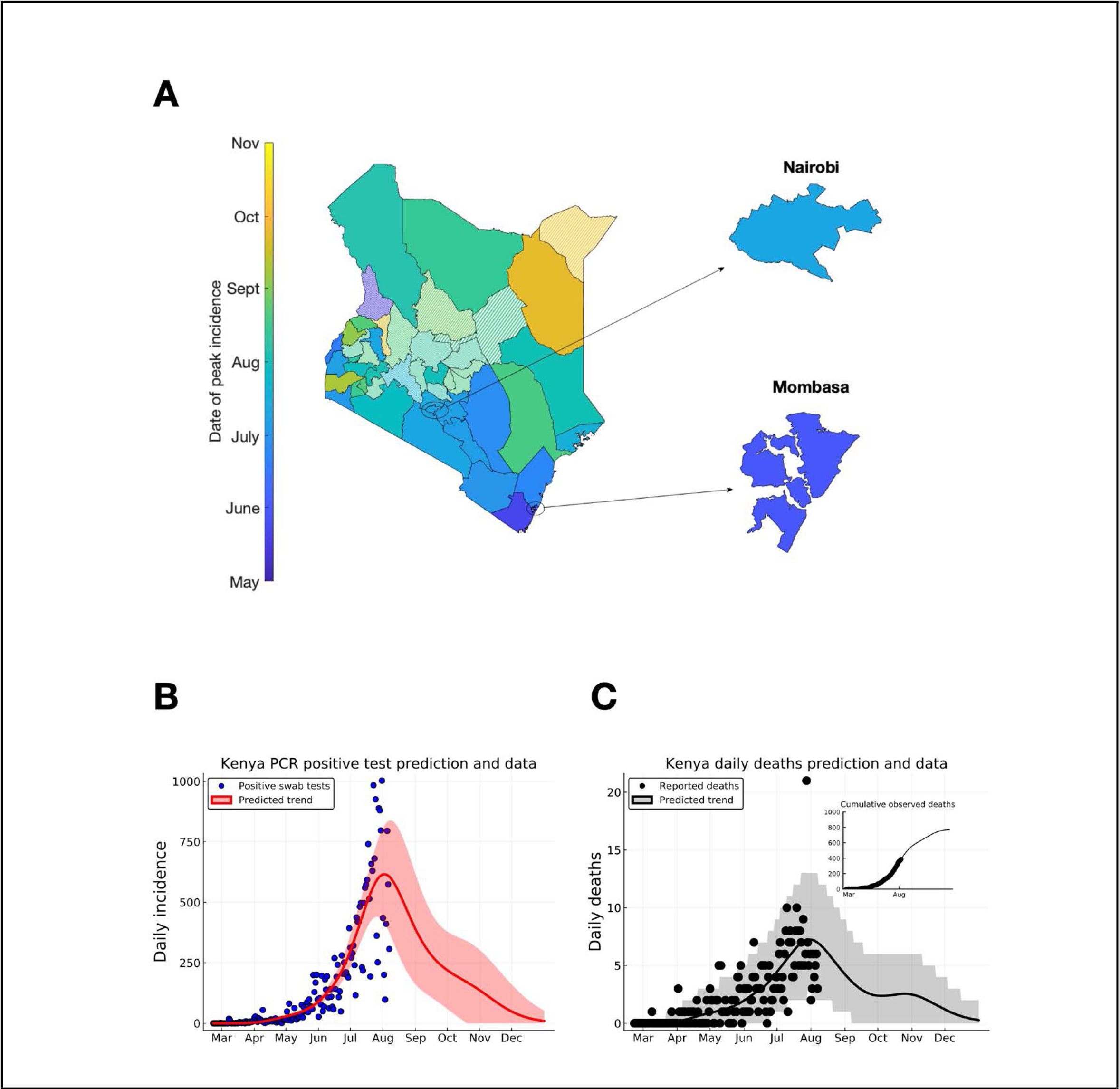
Predicting peak timing of transmission rate by Kenyan county, and forecasting of Kenya-wide PCR positive swab tests and reported deaths. (**A**) Posterior mean for date of transmission rate peak by Kenyan county. Solid shaded counties had sufficient data to infer model parameters with generic weak priors. Candy-striped shaded counties had model parameters inferred with strong priors derived from posterior distributions of parameters for other counties. Inset plots focus on Nairobi and Mombasa cities. (**B**) Kenya total positive swab tests collected by day of sample (*blue dots*) with model prediction of daily positive swab test trend (*red curve*). (**C**) Kenya total reported deaths with a positive swab test (*black dots*), with model prediction of reported death rates (*black curve*). Inset plot indicates cumulative reported deaths with model prediction of cumulative deaths. Dates on (**B**) & (**C**) mark 1st of the month.

Because of the lag between infection and the observability of the infected person (whether by swab PCR test, serology test, or death), we estimate that both daily PCR positive test detections and daily observed deaths attributed to COVID-19 across all of Kenya were peaking in early August 2020. Accessing complete data on hospitalisation rates in Kenya has been challenging. However, sentinel clinical surveillance from county hospitals suggests a modest increase in adult admissions trending towards pre-COVID numbers in June and July 2020 but, on average, lower than observed counts from at a similar period in 2018 and 2019 (*20*). Our prediction is that rates of new PCR positive tests and deaths attributed to COVID-19 will enter a long-tailed decline from August 2020 onwards; that is, the rate of epidemic decline will be less steep than the rapid growth phase as the decrease in the main cities is partially off-set by growth in the rest of Kenya (Fig. 3 B&C). It is likely hospitalisation incidence will follow a similar trend to the infection and death rates. By fitting an IFR_crude_ value for each county in Kenya, we expect that by the end of December 2020 less than 1000 deaths in Kenya will be cumulatively attributed to COVID-19 (Fig. 3C). It should be noted that our projections are based on an assumption that the PCR testing rate in Kenya is either increasing or flat in each county, if the testing rate declines then we expect the rate of PCR positive tests to decline even more steeply than predicted in this study.

Our modelling analysis provides a coherent account of the SARS-CoV-2 pandemic in Kenya. Whilst the full picture of the epidemiology in Kenya will not be established until cause-specific mortality data become available (e.g. from resumption of Demographic Surveillance System and verbal autopsy activities), out model, fitted to three sources of nationwide longitudinal data, suggests that the number of COVID-19 cases and the mortality risk attributable to the SARS-CoV2 epidemic are substantially lower in Kenya than in Europe and the USA. Given that the model suggests 34-41% of the urban population have already been exposed, and that the epidemic has peaked during a period of relatively little restrictions and physical distancing, it seems likely that a significant factor in the epidemic resolving is through population immunity. If this trend continues, then the direct and indirect health and socio-economic consequences of control measures need to be balanced carefully as we obtain more accurate estimates of the consequences of disease.

## Data Availability

The full Kenyan SARS-CoV-2 line list contains sensitive personal information that could potentially allow the identification of individual cases. The analysis performed in this study only required a highly aggregated dataset derived from the Kenyan linelist. Other data used in this paper was openly available. All data is available in the main text or the supplementary materials. The code base underlying the analysis is accessible at the open github repository https://github.com/ojal/KenyaSerology.

## Acknowledgments

We thank all members of Kenya’s county rapid response teams (who collected swab samples), and testing centres (who conducted the laboratory PCR assays) and of Kenya National Blood Transfusion Service Centres. This study is published with the Permission of the Director of the Kenya Medical Research Institute.

## Author contributions

DJN, MJK, JO, SPCB, AA, GMW, JAGS, IMOA, VW, EB, PB Conceptualised and/or designed the study;

JO, SPCB, MJK, IK, RA, CM Formal analysis, methodology, software;

SO, LIOO, CNA Investigation;

BT, AA, EB, KK, PA, MM, RA, WN Administration;

DJN, MJK, EAO, GMW, IMOA, KA Supervision;

MO, EO, LIOO, SO, CNA Data curation;

SPCB, JO, DJN, MJK, PB, EB Writing - Original Draft;

All contributors Writing – Review & Editing

## Supplementary Information

### Materials and Methods

#### Data description

We use three daily time series as sources of data to infer model parameters for each Kenyan county:

- PCR positive swab samples by collection date, (*ObsP*^+^)_(*n,c*_ denoting the number of observed on day *n* in county *c*. The county-specific time series of PCR positive swabs was derived from the Kenya linelist by identifying cases with reported PCR positive swabs in that county by their date of sample collection (or lab confirmation date if collection date was not available). We excluded positive swabs that were traced either at entry into Kenya or due to being a contact of an identified case, so as to focus on Kenyan population surveillance.
- Numbers of sero-positive(*ObsP*^+^)_(*n,c*_ and sero-negative (*ObsS*^−^)_(*n,c*_ blood samples collected from regional centres of the Kenyan National Blood Transfusion Service (KNBS) on day *n* originating from county *c*.
- Daily estimates of relative human mobility *m_n,area_* compared to a baseline of the same date in the previous year (2019) derived from Google mobility trends (*15*).
- Deaths with a positive PCR-confirmed test by date of death for each county, (*X*^+^)_*n*_

The (*ObsP*^+^)_(*n,c*_ time-series ran between 13th March and 10th August, however we removed data from 7th until 10th August because of delay between the sample date and the positive swab test being confirmed and added to the Kenyan linelist. The (*ObsS*^+^)_(*n,c*_ and *ObsS*^−^)_(*n,c*_ data was collected in May and June. Residual blood samples for serology were obtained from regular blood donors attending 4 regional KNBTS centres (Mombasa, Nairobi, Eldoret and Kisumu). The study methodology is fully described in Uyoga et al (*11*).

We assumed that changes in trends in SARS-CoV-2 transmission in Kenya were due to changes in the underlying population mobility. In particular, by changing frequency of indoor congregations. Therefore, we calculated *m_n,area_* as the average change in baseline mobility over the “retail and recreation”, “grocery and pharmacy”, “transit stations”, and, “workplaces” settings (Google defined categories), and also over the week prior to day *n*, in order to average over weekend effects. Due to incomplete data, and the likely bias introduced by using a mobility estimate derived from smartphone users in predicting the mobility of semi-rural populations outside of the major urban conurbations in Kenya, we consider only three areas: Nairobi, Mombasa and the pan-Kenyan aggregate (Fig. S1).

#### Transmission model

The dynamics of transmission were assumed to follow a simple SEIR transmission model with an effective population size parameter (*P_eff_*) (*21*). The *P_eff_* parameter accounted for the effect of population heterogeneity in lowering the proportion of the total population that must become immune for incidence to start decreasing, due to depletion of susceptibles rather than increased social distancing, compared to the prediction of a fully homogeneous model. In homogeneous SEIR models both the early exponential growth rate in incidence and the proportion of the total population that must become immune for incidence rate to start decreasing, e.g. “herd-immunity”, can be determined from the reproductive number (*R*_0_ and the mean durations of latency and infectiousness (*22*). For heterogeneous models of transmission, where potentially different at-risk groups are at different risk of contracting the infectious pathogen and have different infectious potential, determining (*R*_0_ from early growth in incidence aggregated over the different at-risk groups doesn’t give sufficient information to estimate the overall proportion of the population required to become immune before achieving herd immunity. This aspect of heterogeneous models of transmission has been widely investigated, for example, in the context of comparing vaccination coverage thresholds for elimination between uniform and targeted vaccination policies (*23*). In the context of the SARS-CoV-2 pandemic modelling literature, the role of population heterogeneity in lowering the herd-immunity threshold compared to the prediction of a homogeneous population transmission model has again been identified (*24, 25*). In this study, we have taken a phenomenological approach; the effect of heterogeneity in the population was encoded in the effective population parameter *P_eff_*, and this parameter was inferred jointly with (*R*_0_.

The rate of infectious contacts per infectious individual, *γR_t_*, was given as the time-varying *instantaneous* reproductive number (*R_t_*), that is the number of secondary cases per infected individual assuming *both* a fully susceptible population *and* the time-varying mobility rate being fixed at time *t* (*26*), rescaled by the recovery rate from infection (*γ*). The instantaneous reproductive number for a county was assumed to change daily proportional to the change in the mobility rate:

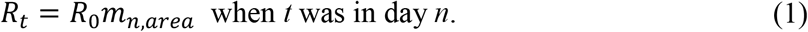

The “area” of the county was Nairobi or Mombasa for those counties, otherwise the Pan-Kenyan mobility estimate was used. The baseline reproductive number *R*_0_ was inferred for each county.

The model dynamics are given as differential equations for susceptible (*S*(*T*)), latently infected (*E*(*t*)), actively infectious (*I*(*t*)), recovered/immune (*R*(*t*)), and, cumulative infections (*C*(*t*)),

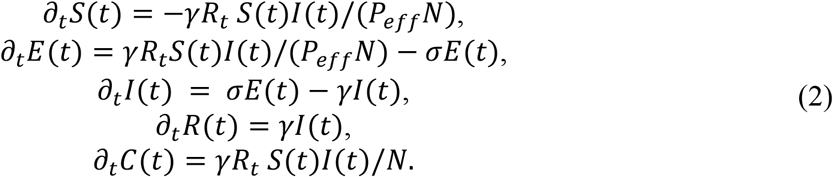

All variables and parameters are described in table S1. The transmission model was initialised on 21st February for each county, 21 days before the first confirmed positive test swab and the first day of available Google mobility data with initial state (*P_eff_N* − *E*(0) − *I*(0), *E*(0), *I*(0),0,0,), where *P_eff_*,*E*(0) and *I*(0) were treated as target parameters for inference.

Equation (2) implies the *effective (instantaneous*) reproductive, that is the instantaneous reproductive ratio where the reduction in susceptibility is also accounted for: 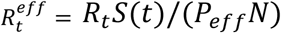.

In total, the transmission model for each county had four unknowns *θ_TM_* = (*R*_0_, *P_eff_*,*E*_0_,*I*(_0_)): the baseline reproductive number (*R*_0_), an effective population size scale (*P_eff_*), the number of latently infected individuals on 21st Feb (*E*(0)), and the number of actively infectious individuals on 21st Feb (*I*(0)).

#### Observation model

The underlying transmission of SARS-CoV-2 is not observed, rather we have access to PCR positive swab tests and serological tests (positive and negative) aggregated by date and county. Therefore, we developed an observation model that connects unobserved daily transmission rates, which depend on the unknown transmission parameters, to a likelihood of the observed test data. The observation model itself had two unknown parameters per county: the mean detection rate per PCR-detectable individual per day by swab testing, and the clustering factor of these detections. By defining a likelihood-based observation model we gained access to modern techniques in Bayesian inference to infer both the unknown parameters of the underlying transmission process and the unknown parameters of the observation process.

The number of people who would test positive, either as PCR positive or as sero-converted, on each day *n* depended on: 1) the rate of new incidence on each day *s < n*, and, 2) the probability that someone who was infected on day *s* is detectable *τ* = *n* − *s* days later. The daily numbers of new incidence (*ι_n_*) on each day *n* predicted by the transmission model was,

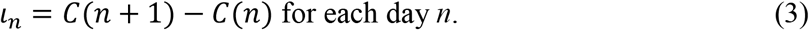

The probability that an infected individual would be determined as having been infected *τ* days after infection by either a PCR test or a serology test was denoted, respectively, *Q_PCR_*(*τ*) and *Q_sero_*(*τ*). The maximum sensitivity of the PCR test in a typical Kenyan setting was part of our inference methodology (see below), therefore we treated *Q_PCR_*(*τ* as being 0 for the first 5 days post-infection (the mean time between infection and symptoms for symptomatic cases (*27*)) and then was fitted to data on diagnostic accuracy given in Zhou et al (*17*), the fitted functional form for PCR-detectability more than 5 days after infection was:

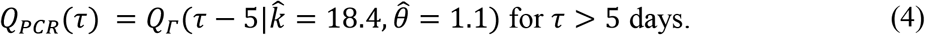

Where *Q_Γ_* was the tail distribution function of a Gamma distribution with fitted shape parameter 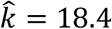 and fitted scale paramater 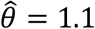. This aligns with Zhou et al that the median period to become PCR undetectable after symptoms was 20 days (CI 17-24 days) (*17*). The lag between symptoms and maximum detectability by serological assays has been reported as 21 days in a recent metastudy of reported diagnostic sensitivities (*28*). We assumed that, given an additional 5 day lag after infection, the sensitivity of the serological assay increased linearly from 0 at infection to a maximum of 82.5% (*11*) over 26 days,

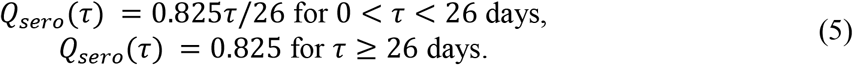

The number of people in the county who would test positive on each day, with a PCR test (*P*^+^)_*n*_, and/or a serology test (*S*^+^)_*n*_, is, therefore,

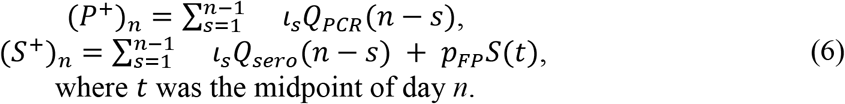

(*p_FP_* was the false positive rate for the serology assay (see table S1). Underlying equation (6) is an assumption that the PCR test is 100% specific to SARS-CoV-2.

The total number of swab samples collected, including negative tests, on each day was not available. The total number of tests performed in coastal counties which were referred to the KWTRP laboratory for PCR was available, and showed considerable variation per day (see Fig S3 for total swab tests per day in Mombasa referred to KWTRP). Moreover, there was variation in the setting at which the swab test was collected, e.g. at hospital, from a walk-up testing facility etc, as the focus of Kenyan public health teams has shifted over the course of the epidemic. Therefore, there was a high degree of variability in the detection probability per PCR-detectable infected individual per day. To account for the high variability in detection, we modelled the distribution of PCR positive swab tests collected on each day using a negative binomial distribution,

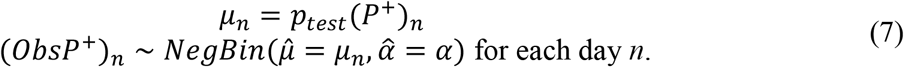

Where *p_test_* was the *mean* detection rate per PCR-detectable, per day. Hence, *μ_n_* is the expected number of positive swab tests that would be sampled on day *n*, given (*P*^+^)_*n*_ PCR-detectable infected individuals, and *α* was the clustering coefficient^1^ for the sampling process: *α* = 0 recovers a Poisson distribution for the number of positive swab tests collected each day, *α* > 0 allowed the model to infer much greater variance in daily positive than expected from a Poisson distribution (*29*). It should be noted that the parameter *p_test_* effectively absorbs swab sample biasing, either for or against selecting individuals who have been exposed to SARS-CoV-2, amongst the swab tested subjects along with the sensitivity of the PCR test as performed in realistic situations.

The number of donor blood samples tested by day of sample collection was available, however, the reported uncertainty in the maximum sensitivity of serology assay was fairly high: the posterior mean sensitivity was 82.5% (credible interval 69.6-91.2%; (*11*)). The posterior uncertainty in the serological sensitivity influenced the confidence the inference method placed on the serological sample data; if the test sensitivity was known to high precision we would treat each days serological samples as a binomial draw from an underlying proportion of seroconverted individuals given by equation (4). Given that the sensitivity of the serological assay was itself an uncertain factor we fitted the posterior uncertainty in the testing sensitivity to a beta distribution: *serological sensitivity* ∽ *Beta*(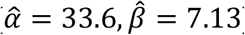). This implied that the appropriate observation model for the number of positive serological samples on day *n* ((*ObsS*^+^)_*n*_), out of the total number of serological samples being collected on day *n*, *N_sero,n_* = (*ObsS*^+^)_*n*_) + (*ObsS*^−^)_*n*_), was a Beta-binomial distribution,

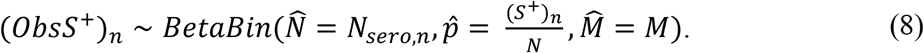

Given an underlying realization of the transmission process the mean number positive serological samples on day *n* is 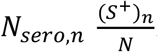. The “total-count” parameter (*30*), 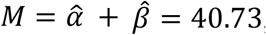, allowed for greater dispersion in the observed seropositive count data than would be allowed by a Binomial model. The unknown parameters of the observation model are *θ_OM_* = (*p_test_*, *α*).

#### Parameter inference for each Kenyan County

The observation model gives the following log-likelihood function for the unknown parameters *θ* = (*θ_TM_*,*θ_OM_*) given the sampling data for a county, ***D*** = {(*ObsS*^+^)_*n*_, (*ObsS*^−^)_*n*_, (*ObsP*^+^)_*n*_}_*n*=1,2,…_

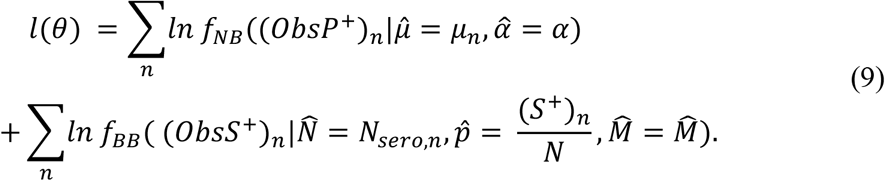

Where *f_NB_* and *f_BB_* are, respectively, the probability mass functions for the negative binomial and beta-binomial distributions, as described in the <u>observation model</u> subsection. The first day where samples were included in the log-likelihood calculation was 12th April, due to testing being even more irregular before that date (Fig. S2), and the last day was 6th August, due to right-censoring of samples collected on 7th-10th August appearing in the linelist dated 10th August.

We divided the Kenyan counties into two groups: those counties with and without serology samples. The availability of serology data was highly important for this study because without serology data, the detection rate was not identifiable jointly with *P_eff_* and the initial conditions using weak priors. Therefore, we inferred unknown parameters in the counties with serology data using weak priors (priors described below), whereas for counties without serology data we used strong priors based on the posterior distribution of the counties that had serology data (method described below). There were two exceptions to the county grouping: Kajiado county, which has no attributed serology samples but a substantial number of PCR-positives, and was added to group of counties fitted using weak priors, and West Pokot county, which had some serology data but small numbers of PCR-positives, and was added to the group using strong priors.

The prior distributions for *R*_0_, *P_eff_*, and *α* were the same for each county using weak priors: 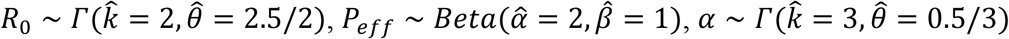. These priors reflect weak confidence that: 1) the fundamental *R*_0_ value in Kenya would be similar to that seen in the early Chinese epidemic, 2) the most *apriori* likely value of *P_eff_* was 1, thus recovering the standard homogeneous SEIR model, and 3) that the daily detected PCR-positive swab data would be moderately overdispersed compared to a Poisson distribution. The prior distributions for *p_test_* and the initial numbers of infected *E*(0) + *I*(0) in counties with weak priors differed by county:

- **Nairobi**. 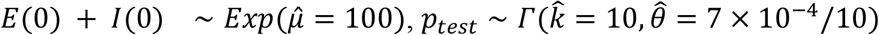
- **Mombasa**. 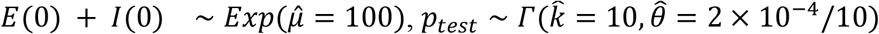
- **Kajiado, Kiambu, Kilifi, Kisumu, Machakos, Migori, Uasin Gishu, Vihiga counties**. 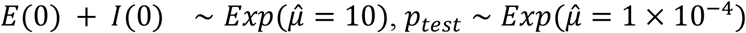
- **Bungoma, Busia, Garissa, Homa Bay, Kericho, Kisii, Kitui, Kwale, Lamu, Makueni, Marsabit, Migori, Narok, Nyamira, Nyeri, Siaya, Taita Taveta, Tana River, Trans Nzoia, Turkana, Wajir counties**. 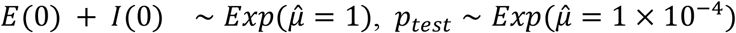

The choice of weak priors for the counties with serology data was informed by estimates of urban density, and reflect our belief that the epidemic in Kenya was seeded by international travel to both Nairobi and Mombasa international airports and that there was *apriori* likely to be a higher number of infected individuals in Nairobi and Mombasa on February 21st. Our prior belief about mean detection rate was informed by openly available testing rate data for Kenya (*31*) (Fig. S3), which was likely to be mainly concentrated around Nairobi, available data on testing rate in Mombasa (Fig. S2) and a weak belief that detection elsewhere would be about half as effective as in Mombasa.

The prior distributions for all parameters for counties without serology data were much stronger, reflecting that in the absence of significant amounts of data, we expected these counties to have similar parameters to other Kenyan counties. To create the strong priors we pooled the posterior parameter draws of exemplar counties, and then projected this multivariate empirical distribution onto a collection of independent marginal distributions by applying maximum likelihood to each set of parameters (each parameter was assumed to have a Gamma marginal distribution except *P_eff_*, which was assumed to have a Beta marginal distribution. Therefore, inference in the counties without serology penalised against drawing parameters that were outside the univariate credible intervals of their exemplar county. The exemplars used were:

- **Isiolo and Nakuru counties**. Exemplar counties were Kajiado and Machakos.
- **Baringo, Bomet, Elgeyo Marakwet, Embu, Kakamega, Kirinyaga, Laikipia, Mandera, Meru, Murang’a, Nandi, Nyandarua, Samburu and Tharaka Nithi, West Pokot counties**. Exemplar counties were Homa bay, Tana River, Turkana, and Wajir counties.

The choice of exemplar counties was based on Kajiado and Machakos containing a mixture of urban and rural dwelling broadly similar to Isiolo and Nakuru counties. Whereas, the other Kenyan counties lacking serology data were predominantly rural. Therefore, we used a mixture of four rural exemplar counties distributed over the country.

We used Hamiltonian MCMC (*32*) to perform Bayesian inference by drawing 10,000 samples from the posterior distribution,

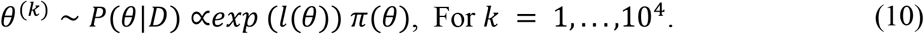

for each county using the NUTS-HMC sampler implemented by the Julia language package *dynamicHMC.jl*. Solving the likelihood function for a proposed value of involved solving the ODE system (2), we used the highly performant DifferentialEquations.jl package for ODE solutions (*33*). The HMC method required a log-likelihood gradient, *∇_θ_l*, which, for our use-case of a small ODE system with a low number of parameters, was most efficiently supplied by forward-mode automatic differentiation (*34*) implemented by the package *ForwardDiff.jl*.

The MCMC chain converged for each county (all MCMC chains and MCMC diagnostics can be accessed through the linked open code repository). The posterior mean (and 95% Cis) for each parameter, as well as date of maximum infection rate and estimated county-specific IFR, can be found in supplementary Data S1. West Pokot county (1 positive PCR-confirmed test and 2/17 positive serology samples) was an outlier in the analysis in that the inference converged on a posterior mean estimate of *R*_0_ which was less than one.

#### Nowcasting and Forecasting for Kenyan counties

The parameter draws from the posterior distribution defined the uncertainty in our model nowcasts and forecasts for each county, since the underlying transmission model was deterministic. Therefore, posterior distributions for epidemic quantities were created by simulating the epidemic for each *θ*^(*k*)^, *k* = 1,2,…, 10^4^ posterior draws, for example the draws from the distribution of day of peak infection rate were,

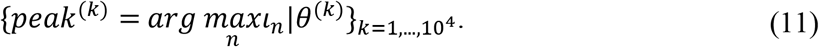

The central estimate is the posterior mean, 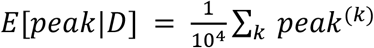. and the 95% credible intervals were the 2.5% and 97.5% empirical quantiles of the draws from the day of peak. This approach was used through-out the main document. It should be noted that for the daily positive PCR tests we showed both the posterior mean and CIs for the mean daily detection rate of new PCR-postive swab tests, *E*[*μ_n_*|*D*], and the posterior prediction interval, *E*[*ObsP*^+^)_*n*_|*Dμ_n_*|*D*] (see equation (7)). The two quantities have the same posterior average, but (*ObsP*^+^)_*n*_|*D* has substantially higher variance since it includes both uncertainty in the parameter estimates *and* the fundamental variance in the observed daily detection of positive swab tests.

#### Inferring a crude infection fatality ratio

The commonly used infection fatality ratio (IFR) by age estimates from Verity et al (*19*), weighted by the Kenyan population distribution given by the 2019 census, implied a basic IFR prediction in Kenya of *IFR_verity_* = 0.264%; that is 264 deaths per 10,000 infections. This assumed an uniform attack rate across age-groups in Kenya.

We used the posterior predictions of the underlying daily infections in Kenya counties to infer a crude infection fatality ratio (*IFR_crude_*) for each Kenyan county. The lag between infection and death, for those infected individuals who die, was defined as the convolution between three time duration distributions:

1. The duration of time between infection and symptoms (days), which we assumed was distributed 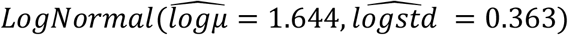 (*27*)
2. The duration of time between initial symptoms and severe symptoms (days), sufficient to seek hospitalisation, which we assumed was distributed *U*(1,5)(*35*).
3. The duration of time between severe symptoms and death estimated from UK hospital data (*36*). This was an empirical distribution with mass function *p_HD_*

We discretized the first two distributions to give probability mass functions *f_IS_* for the number of days between infection and symptoms, and *p_SH_* for the number of days between symptom onset and severe symptom onset. The probability mass function for the (discrete) number of days between infection and death, for those who died, *p_ID_*, was given as a discrete convolution over these probability mass functions:

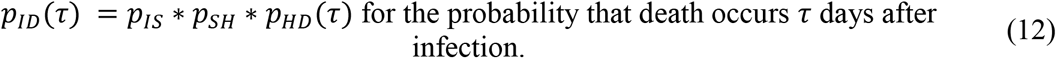

The most likely lag between infection and death was 14 days, however, the distribution was fairly heavy-tailed with mean lag between infection and death 19 days (Fig. S4).

We assumed that the number of deaths observed each day were Poisson distributed, and accounted for the lag between infection and death,

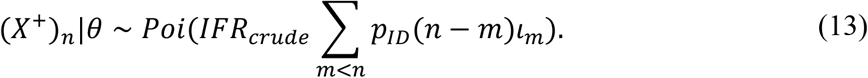

The conditionality on *θ* is given to emphasise that the number of infections per day depended on the unknown parameters for the county. Using 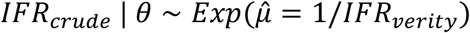 as a weak prior for each of the county specific estimates of the crude IFR observed in Kenya, conditional on a realization of *θ*, we found that the posterior mean estimator took a simple form (see <u>Posterior distribution of crude infection fatality ratio</u> in supplementary text below):

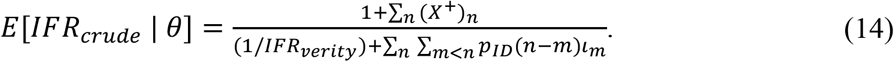

Similar to equation (11), we approximated the full posterior distribution of *IFR_crude_*|(*X*^+^)_1_,…, (*X*^+^)_*N*_, *D* using a set of draws: {*IFR*^(*k*)^ = *E*[*IFR_crude_*|*θ*^(*k*)^]} _= 1,…,10_^4^ The posterior mean estimator and credible intervals for the county-specific*IFR_crude_* values were calculated from this set of draws.

#### Model refinement to account for increased test rate at the capital

The publicly available *ourworldindata.org* coronavirus dataset (*31*) showed that the daily number of tests in Kenya increased from 29th March at an approximate rate of 1.6% per day relative to the overall mean tests per day (mean 2482 tests per day in Kenya between 29th March - 3rd August; Fig. S3). This increase in testing was not reflected in increased testing at KWTRP testing laboratory (Fig. S2), therefore, we hypothesised that the increased rate of testing in Kenya was being driven by increased testing in Nairobi and the surrounding counties of Kajiado, Kiambu and Machakos. To test this hypothesis we considered an alternate model for detection where *p_test_* is calendar day dependent:

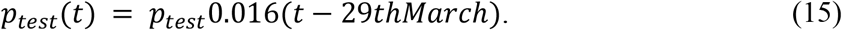

The baseline mean detection rate *p_test_* remained the target of inference. The same period for inference (12th April - 6th August) was used.

Incorporating the increased testing rate into the inference model improved the model information criteria relative to the basic hypothesis that mean detection rate had been constant since 12th April. This was observed using two different model information scores: the popular in-sample Bayesian model fit score, Deviance information criterion (DIC; (*16*)) defined as *DIC* = −*E*[*l*(*θ*)|*D*] + 2*var*[*l*(*θ*)|*D*], and the out-of-sample log predictive density (LPD; (*16*)) for deaths. The LPD for a model was defined as,

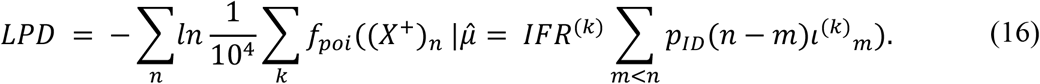

The LPD measure is the sum log probability of observing the death data for the county, *f_poi_*(*x*|*μ*) was the probability mass function for a Poisson distribution with mean *μ*, the emphasises that the number of infections on day *m* depends on the *k*^th^ draw of the parameters. Although the IFR was optimised to fit the death data, we also wish to emphasise that the deaths were not used to infer other model parameters; this justified our description of LPD as an out-of-sample metric.

Both the DIC and LPD model information criteria for Nairobi were significantly improved by including the increasing detection rate in the model [equation (16)]: Δ*DIC* = 10.8 and Δ*LPD* = 12.4. Therefore, for Nairobi, and its neighbouring counties with observed large numbers of positive tests: Kajiado, Kiambu, and Machakos, we used the increasing detection rate model in the main manuscript results.

#### Model validation: Posterior predictive P-values

We validated the overall fit of the best performing model (increasing detection rates over the inference period for Kiambu, Kajiado, Machakos and Nairobi counties; flat detection rates for other counties) using posterior predictive checking of the LPD value (*30*). Posterior predictive checking is a Bayesian model checking analogy to classical statistical chi-squared tests or G-tests. For each county, after inferring a posterior distribution for the unknown model parameters and the county specific IFR estimate, we sampled 1000 *replicated* death data time series, 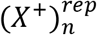, each of which had their own LPD value. The observed distribution of LPD values represented the expected distribution of log-predictive densities *if the data was really generated from the model* {(LPD^rep^)_*k*_ *k* = 1,…,1000}. The posterior predictive P-value for the model is defined as 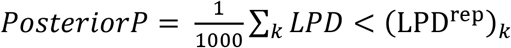; that is the observed proportion of replicated LPD values from the model that were greater than the observed LPD value of the true data.

The average value for the posterior P-value over the counties was 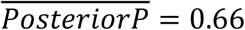 (minimum-maximum spread: 0.12-0.85, Data S1), therefore, the model was typically *more accurate* at predicting the daily number of deaths actually observed than replicated data. This might be caused by deaths occurring with lower variance than predicted by a Poisson distribution.

### Supplementary Text

#### Notation for distributions used in this study

In this study, we have used a number of parameter symbols that are also the most commonly used symbols for various common parametric distributions. Moreover, these parametric distributions are used in the underlying analysis frequently with their distribution parameters defined as functions of underlying transmission model states. To reduce misunderstanding reserve symbols with “hats” as referring to the parameters of a parametric distribution and use “=” to refer to the value of the parameter. Find below the choice of parametrization for the parametric distributions used in the study:

- Exponential distribution. 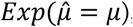, with mean 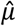
- Gamma distribution. 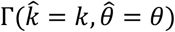, with shape parameter 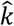 and scale parameter 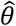
- Negative binomial distribution. 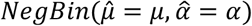, with mean 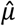 and clustering factor (inverse dispersion parameter) 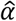
- Beta distribution. 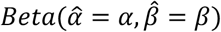, with shape parameters 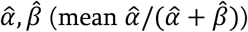.
- Beta-binomial distribution. 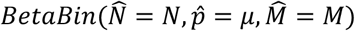, with number of draws 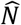, marginal probability per draw 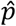, and “total-count” 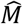 (see (*30*) for details of this slightly unusual parameterization).
- Poisson distribution. 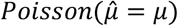, with mean 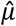.

#### Posterior distribution of crude infection fatality ratio

Here we give details of the posterior distribution of *IFR_crude_*. It is well known that the exponential distribution is a conjugate prior to the mean of a Poisson distribution; that is that given a model where data 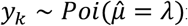 for *k* = 1,…, *N*, and a prior 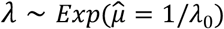, then the posterior distribution of λ is gamma distributed with an analytical solution: 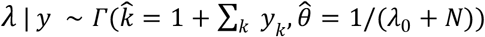. We demonstrate that the posterior crude infection fatality ratio, *conditional on some fixed value of the parameters θ*, *IFR_crude_*|(*X*^+^_1_),…,(*X*^+^)_*N*_,*θ*, and with the prior distribution 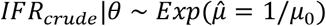 is also gamma distributed. This holds despite the mean number of deaths changing daily, and the posterior distribution only depends on the total number of observed deaths and the cumulative number of time delayed infections. First, we define the time delayed infections on each day using the lag distribution *p_ID_* [equation (12)], 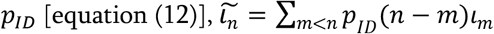. We define the total time delayed infections, 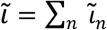, and the total deaths, 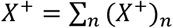.

Then, after cancelling terms, the posterior distribution of *IFR_crude_*|*θ* given the daily deaths is,

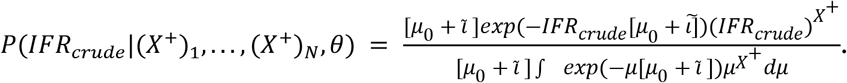

By identifying the denominator as the (*X*^+^)^th^ moment of an exponential distribution we arrive at,

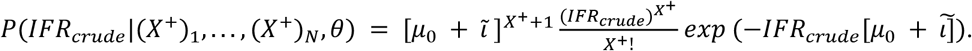

The right hand side of this expression is the density function of a 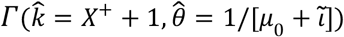 distribution, that only depends on the total deaths and time-delayed infections. Therefore, the posterior mean estimator for the crude infection fatality ratio conditional on the death data and a fixed value of *θ* is:

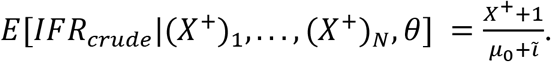

**Fig S1:**
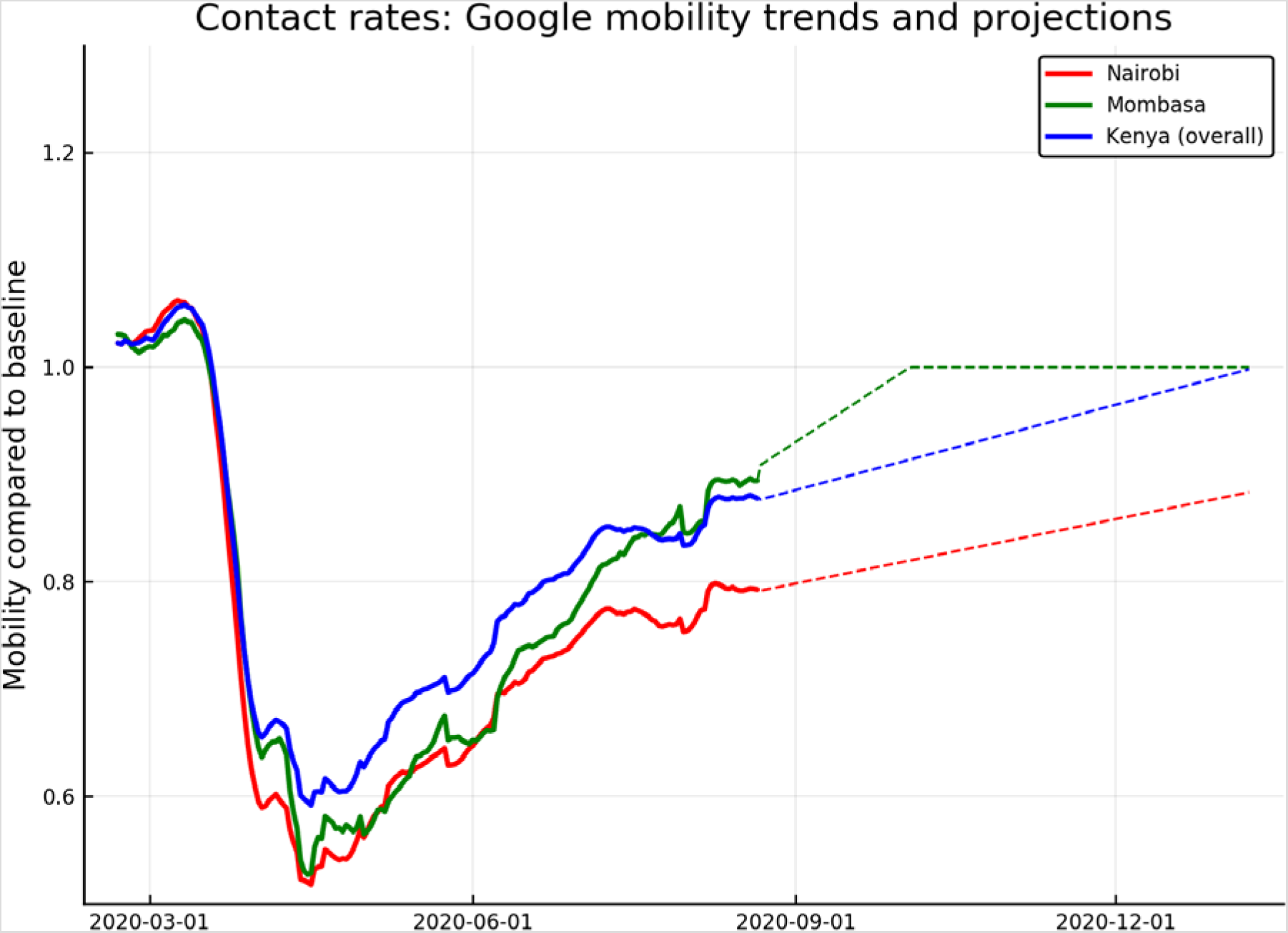
Google mobility trends. The mobility trends used in this study. The curves show a 7-day moving average of the mean relative mobility in the, Google defined setting categories, “retail and recreation”, “grocery and pharmacy”, “transit stations”, and, “workplaces” for Nairobi (*red curve*), Mombasa (*green curve*), and the overall Kenyan trend (*blue curve*).

**Fig S2:**
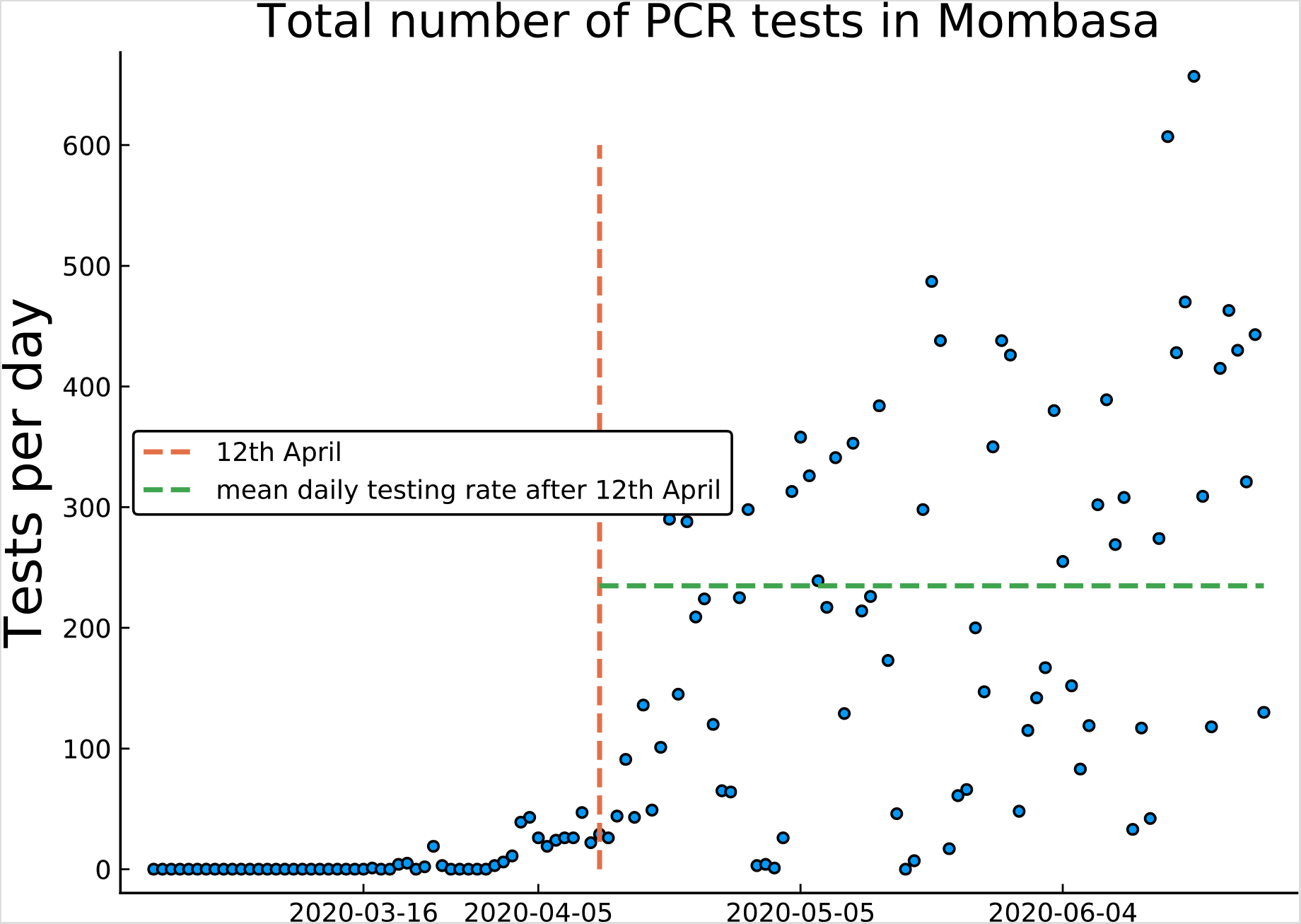
Daily number of swab test kits analysed at KWTRP testing centre for samples collected in Mombasa county. Total number of tests laboratory confirm (positive or negative) (*blue dots*) from 21^st^ Feb until 27^th^ June (data on confirmed negative swab tests was not available for after this date, Data S5). 12^th^ April is shown as a red dashed line; samples before this date were not used in inference because of low testing rates across Kenya. The mean number of tests collected per day in Mombasa after 12^th^ April was 223 (*green dashed line*).

**Fig S3:**
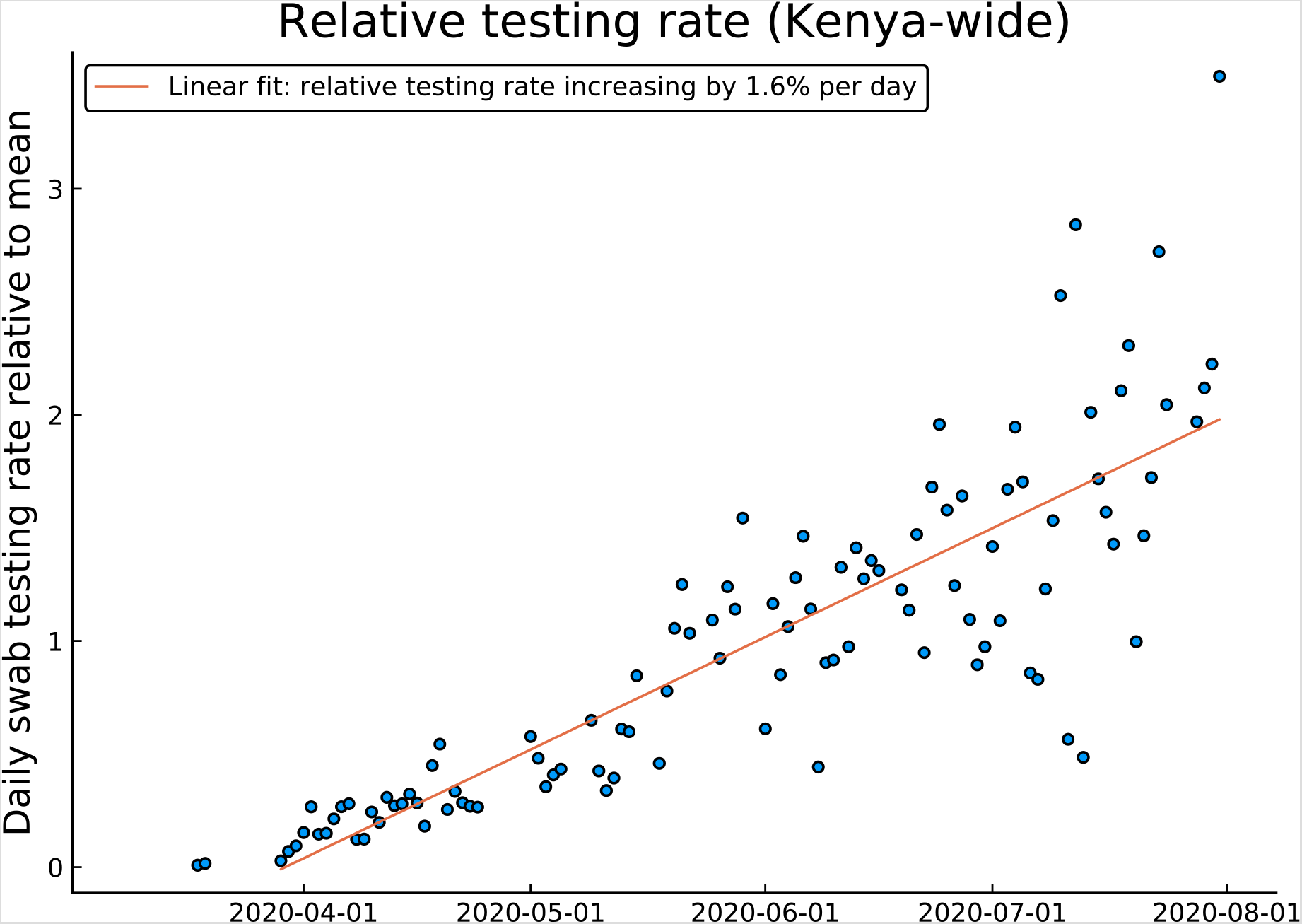
Trend in daily testing overall Kenya since 29th March. The number of tests collected each day reported as a Kenya-wide statistic relative to a mean testing rate of 2482 tests per day across Kenya (*blue* dots; data available from an open source repository (*31*), Data S6). Testing rate increases approximately linearly (*red line*) with a trend of 1.6% increase in testing relative to the mean (40 additional tests per day).

**Fig S4:**
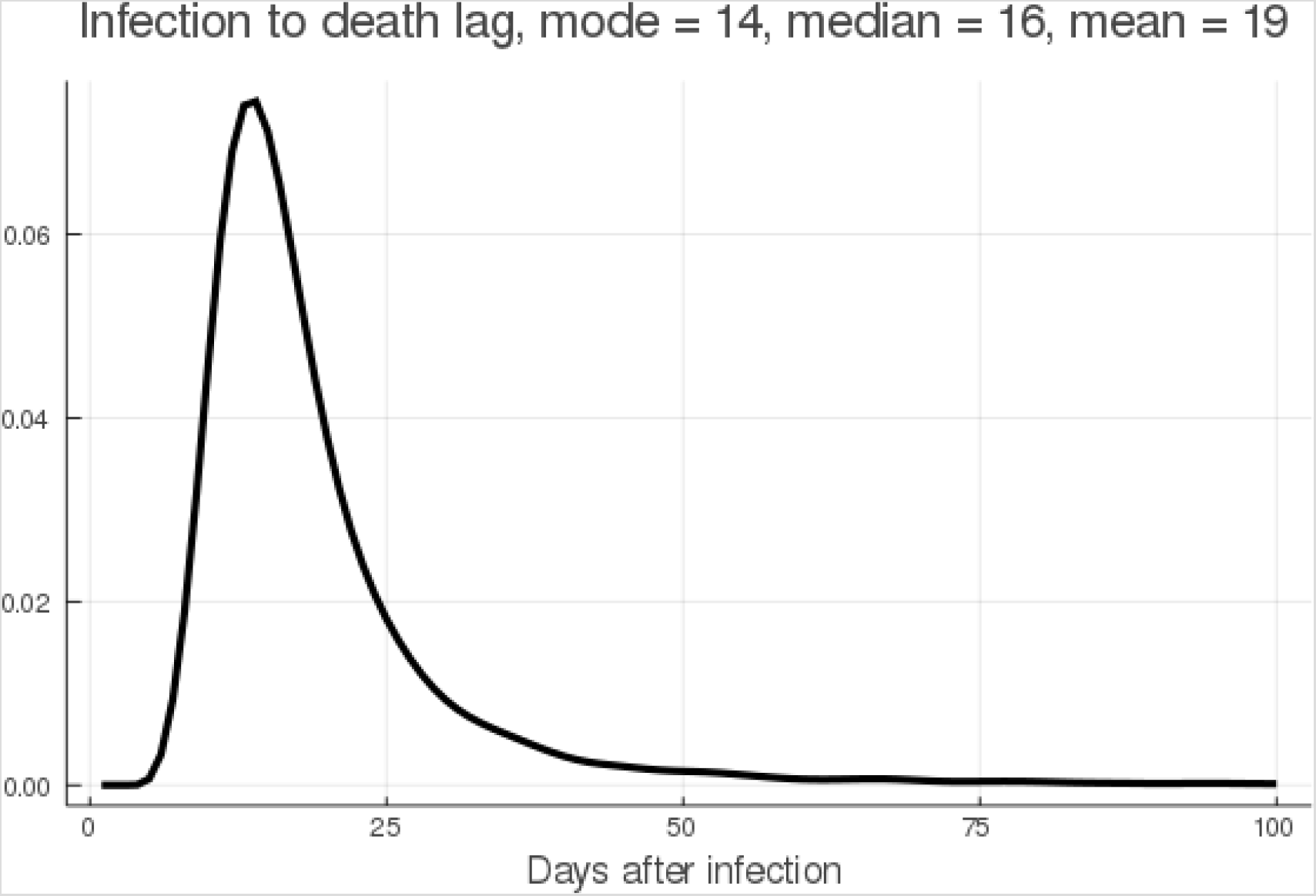
Distribution of time between infection and death for those infected who die. The empirical distribution of time between infection and death, conditional on a death outcome, derived as a convolution over published distributions of durations between infection and symptoms, symptoms and severe symptoms, and severe symptoms and death.

**Table S1:**
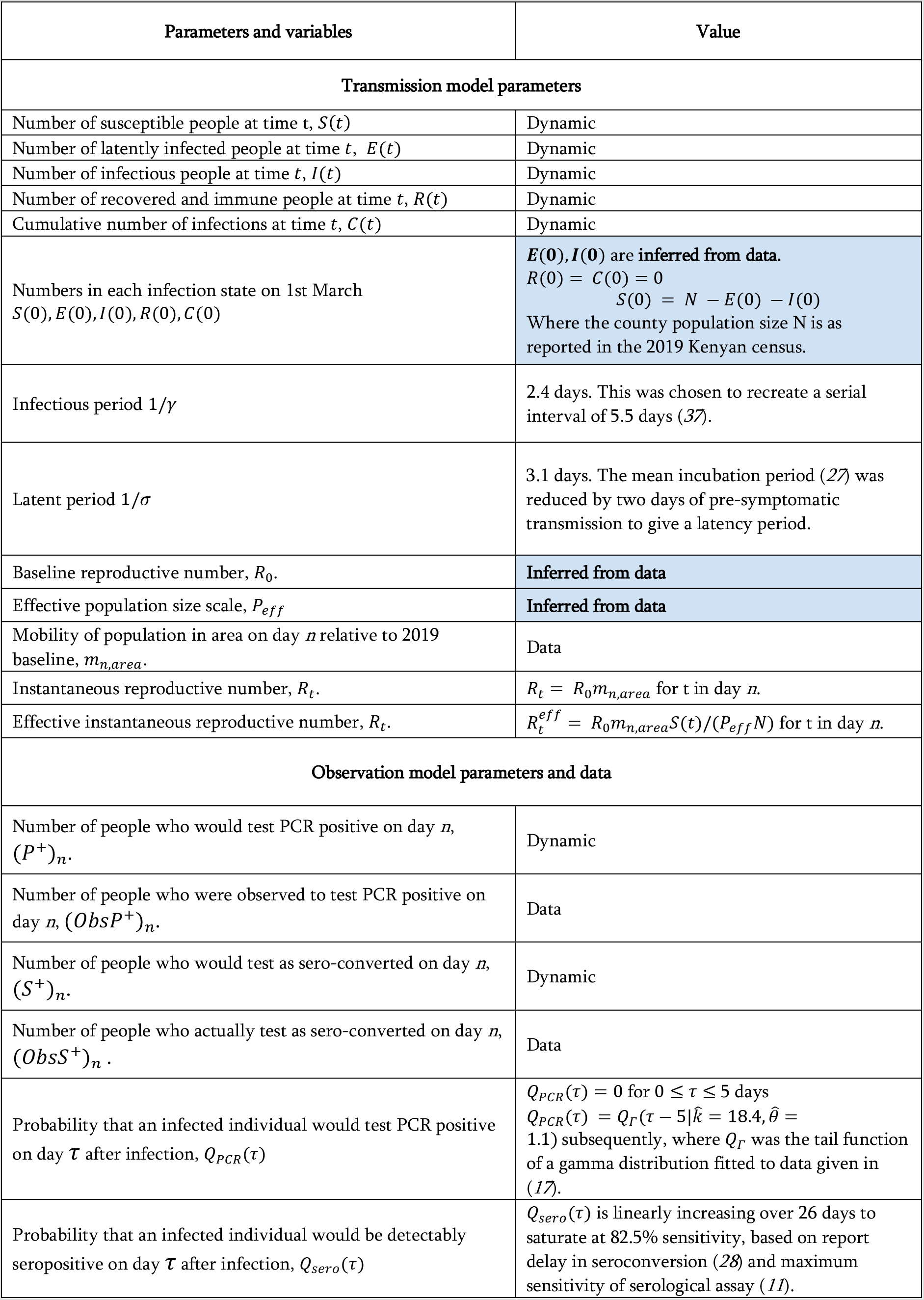
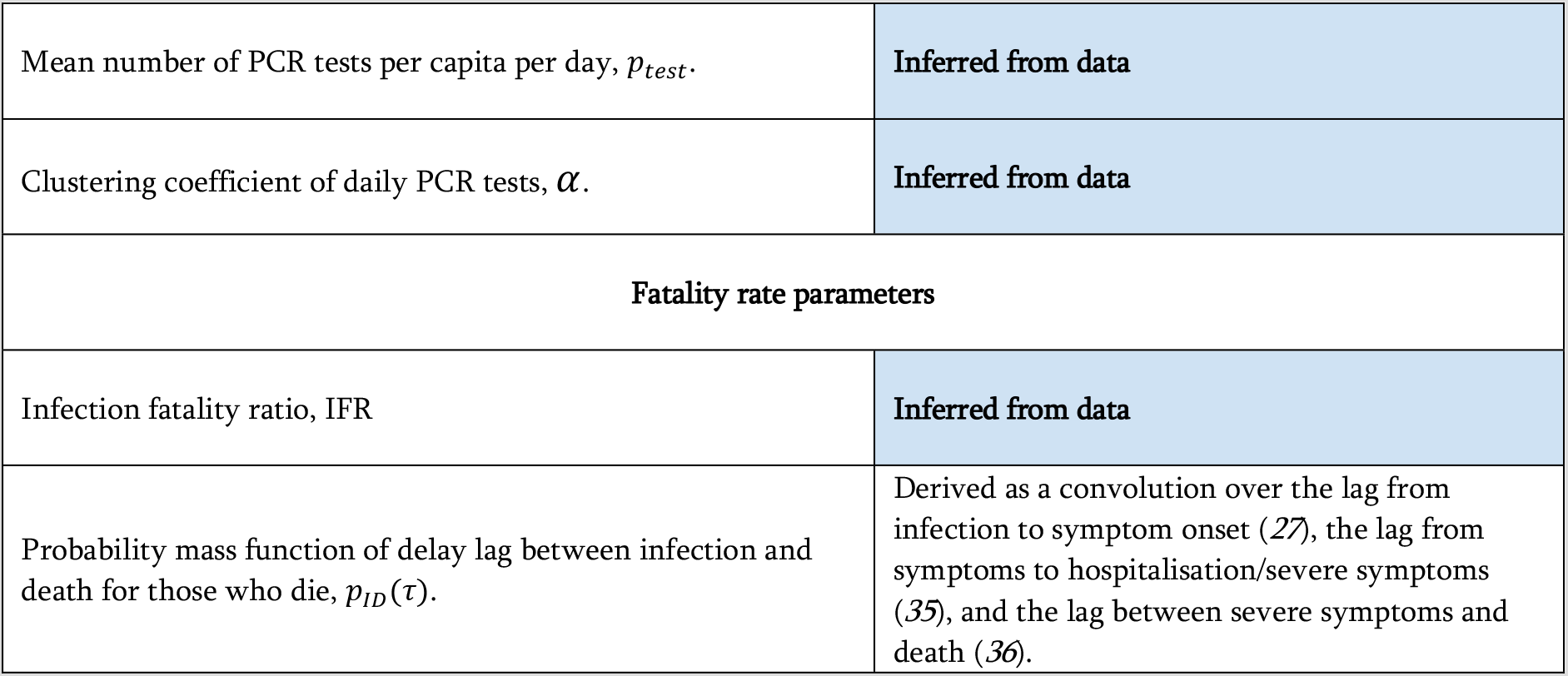
Dynamic and observational model variables and parameters. “Dynamic”, means that the variable was an output of the transmission and observation model for the county.(*37*)

**All datafiles described below can be found in the opendatacsvs/ folder on the github repository**.

#### Data S1. (separate file)

**Inferred parameters and posterior predictive P-values for each Kenyan county**. The posterior mean and 95% credible intervals for each model parameter (both transmission and observation models), with posterior mean and 95% credible intervals for the peak day of infection rate and county-specific infection fatality ratio. Final column is the proportion of synthetic replicated death time series with greater log-predictive density score than the observed death time series for that county (the posterior predictive P-value for the model).

#### Data S2. (separate file)

**The number of positive PCR-confirmed swab tests for each county by date of sample collection (21^st^ Feb to 6^th^ August)**.

#### Data S3. (separate file)

**The number of positive and negative serological results for each county by date of sample collection (21^st^ Feb to 6^th^ August)**.

#### Data S4. (separate file)

**The number of deaths with a PCR-confirmed swab test for each county by recorded date of death (21^st^ Feb to 6^th^ August)**.

#### Data S5. (separate file)

**Total number of swab samples collected in Mombasa county, and analyzed at Kemri-Wellcome Research Program testing centre (21^st^ Feb – 27^th^ June)**.

#### Data S6. (separate file)

**Summary data of Kenyan epidemic, including reported total number of test performed**.

## Footnotes

1 Here the clustering coefficient is the inverse of the dispersion parameter *k*, a common alternative parameterisation of the negative binomial distribution.

